# Elevated mucosal antibody responses against SARS-CoV-2 are correlated with lower viral load and faster decrease in systemic COVID-19 symptoms

**DOI:** 10.1101/2021.02.02.21250910

**Authors:** Janeri Fröberg, Joshua Gillard, Ria Philipsen, Kjerstin Lanke, Joyce Rust, Diana van Tuijl, Teun Bousema, Elles Simonetti, Christa E. van der Gaast – de Jongh, Mariska Bos, Frank J. van Kuppeveld, Berend-Jan Bosch, Marrigje Nabuurs-Franssen, Nannet van der Geest-Blankert, Charlotte van Daal, Martijn A. Huynen, Marien I. de Jonge, Dimitri A. Diavatopoulos

## Abstract

Mucosal antibodies play a key role in protection against SARS-CoV-2 exposure, but their role during primary infection is not well understood. We assessed mucosal antibody responses during primary infection with SARS-CoV-2 and examined their relationship with viral load and clinical symptoms. Elevated mucosal IgM was associated with lower viral load. RBD and viral spike protein-specific mucosal antibodies were correlated with decreases in systemic symptoms, while older age was associated with an increase in respiratory symptoms. Up to 42% of household contacts developed SARS-CoV-2-specific mucosal antibodies, including children, indicating high transmission rates within households in which children might play an important role.

## INTRODUCTION

Transmission within households is an important contributor to the spread of SARS-CoV-2, as close contact within households facilitates early-onset pre-symptomatic transmission of the virus.^1-3^ The rapid spread of SARS-CoV-2 in populations is attributed to several aspects of the virus, i.e. route of transmission via respiratory droplets, rapid viral replication and shedding from the upper-respiratory tract^4^, early infectiousness with a peak viral load before onset of symptoms^5,6^, and a high frequency of mild and asymptomatic infections^6-10^. These aspects make controlling the outbreak difficult, as current control strategies are primarily dependent on symptomatic case detection^11,12^. Additionally, they indicate a high probability that pre-symptomatic carriers are important drivers of community-based transmission of the virus^11,13^. The role of children in transmission remains a highly debated subject, as children often develop asymptomatic or atypical disease which makes them prone to underdiagnosis^3,9,14^. A better understanding of the virology, immunology and clinical symptoms of mild SARS-CoV-2 infections in a community setting is therefore essential to track the spread of SARS-CoV-2 in the general population, and to inform intervention measures.

Antibodies play a crucial role in the protection against viral re-infection by neutralizing the virus upon re-entry. SARS-CoV-2 enters human cells by binding to the ACE2 receptor with the receptor binding domain (RBD) of the Spike (S) protein. Serological studies have shown that antibodies directed against these antigens are capable of hindering SARS-CoV-2 infection, and vaccines targeting the S protein have been shown to be efficacious ^4,6,15,16^. Infection with SARS-CoV-2 also induces humoral responses against the highly immunogenic nucleocapsid (N) protein. The N protein of SARS-CoV-2 shares approximately 80% of its amino acid sequence with SARS-CoV-1 and other seasonal coronaviruses^17^. Therefore, pre-existing immunity against the N-protein may play a protective role during infection^17,18^. Studies investigating antibody response dynamics in mild cases have demonstrated the development of serum antibodies against SARS-CoV-2 approximately 10-15 days post symptom onset^4,19^. Relevance of serum antibodies to protection of severe disease is however equivocal, as some studies found that the antibody response in mild infections is transient^20,21^, while others reported robust and long-lasting antibody responses following mild infections^22,23^.

An understudied aspect of the immune response to SARS-CoV-2 infection is the magnitude and kinetics of the mucosal antibody response. Studies of other coronaviruses conducted with controlled laboratory exposure have shown that mucosal antibodies play a key role in the reduction of viral load and may contribute to protection following re-exposure^24,25^. However, the vast majority of antibody studies on SARS-CoV-2 in humans thus far have focused on serum measurements, with relatively little attention to mucosal antibodies. To obtain a comprehensive view on the role of mucosal immune responses in mild SARS-CoV-2 infection, we performed a prospective, observational household contact study. We assessed the timing, magnitude and complexity of mucosal antibody responses against SARS-CoV-2 antigens and examined their associations with viral load and COVID-19 related symptom development. Additionally, serological and mucosal antibody measurements were used to identify additional cases among close household contacts that were tested negative by PCR at study start. We observed the strongest increases in mucosal antibodies for IgM and IgG directed against S and RBD early after symptom onset, with elevated mucosal IgM levels associated with lower viral load. Increased RBD and S-specific mucosal antibodies correlated with decreases in systemic symptoms over the study period, while older age was associated with an increase in respiratory symptoms. Finally, we demonstrate that up to 42% of participating household contacts develop antibodies to SARS-CoV-2, including children, suggesting high transmission among household contacts. Child contacts were infected at a similar rate as adult contacts with similar viral load, but developed less symptoms compared to adults. Therefore the role of children in transmission might be underestimated.

## RESULTS

### Cohort description and study design

The recruitment strategy for inclusion of households focused on healthcare workers with a PCR-confirmed infection who were in home isolation (index cases) with at least two participating household members. Between 26 March 2020 and 15 April 2020, we enrolled 50 index cases and 137 household members **(Figures 1a and b)**. Home visits were planned to collect naso and oropharyngeal swab samples and nasal mucosal lining fluids (MLF) on the day of study enrolment (D0). In addition, MLFs were self-sampled on three subsequent timepoints and a serum sample was collected on the last day of the study. Index cases were asked to report their first day of symptoms, and all volunteers were asked to complete a daily symptom survey in order to monitor disease progression **(Figure 1c)**. The analysis of mucosal antibody responses was compared with viral load, serological and symptom data. The general characteristics of study participants are shown in **<u>Table S1</u>**. Index cases were mostly female (76%) which is reflective of the gender amongst healthcare workers, with a median age of 46 (IQR: 37-54), while household members were mostly male (61%) and younger, with a median age of 21 (IQR:13-46). At inclusion, 46 (92%) of the index cases and 48 (35%) of the household members were PCR positive.

**Figure 1.**
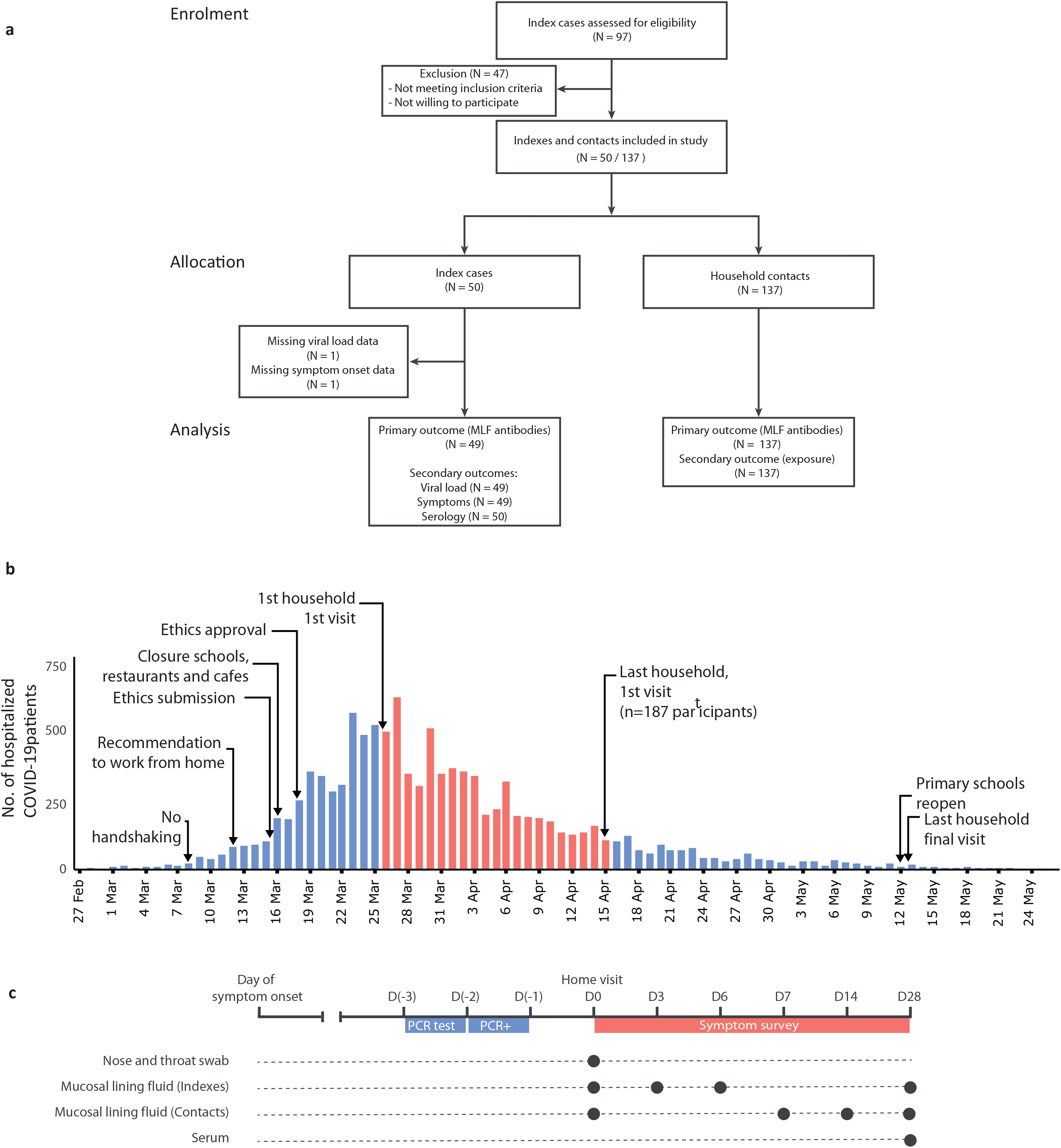
Flow diagram and study procedures. **a)** Flow diagram describing the recruitment of households, sample sizes, and study outcomes. We initially contacted 97 index cases that were tested positive for SARS-CoV-2. After exclusion of cases that did not meet the inclusion criteria or did not consent, 50 index cases and their household contacts (N=137)were recruited. Mucosal lining fluid (MLF) antibodies were analysed as a primary outcome in both indexes and household contacts. Secondary analyses (correlation of MLF antibodies with viral load and symptoms, serology, estimation of SARS-CoV-2 exposure) were performed. **b)** Study timeline, with respect to the number of hospitalizations due to COVID-19 over time and COVID-19 control measures in the Netherlands60,61. The first home visit was conducted at the peak of hospitalizations at March 26, and the last visit was one day after the reopening of primary schools, at May 13. **c)** Overview of the study design and measurements. Home visits were initiated after the index was tested positive for SARS-CoV-2 by PCR, to collect naso- and oropharyngeal swabs for viral load determination as well as nasal MLF samples. Subsequent MLF samples were collected and stored by the participants, who also completed a daily symptom survey. At the end of the study, blood samples were collected for serological analyses.

### Serum antibody responses against SARS-CoV-2 in index cases and their household members

The magnitude of antibody responses was measured in serum and MLF using a fluorescent-bead-based multiplex immunoassay to quantify IgG, IgA and IgM isotypes specific for S-N- and RBD-antigens. We divided the log2-transformed antibody levels by the mean of control samples collected in the pre-SARS-CoV-2 period (n=32 for serum, and n=17 for MLF), to create a normalized value (relative ratio) that reflects the signal over background measurements.

Serum antibody responses, measured at the end of the study, were significantly higher in index cases than in controls for all antigens and isotypes **<u>(Figure S1)</u>**. In order to evaluate differences between antibody isotype and antigen specificities, index cases were classified as seroconverted for each antigen and isotype, based on the difference between their sample and the negative control samples. By the end of the study, 96% of index cases and 53.28% of household members (contacts) seroconverted based on at least one antibody measurement, and the highest seroconversion rate was found for the S-protein **(Figure 2a)**. Nearly all index cases (96%) seroconverted for S-protein IgG and IgA, while the seroconversion rate for RBD and N-protein was substantially lower or even absent (50 and 70% for IgG and 2 and 0% for IgA, respectively). Similar patterns were found in the household contacts. In order to compare local (respiratory) versus systemic humoral immune responses, we correlated serum and MLF measurements taken at the end of the study. All correlations were positive and statistically significant, and were strongest for the IgM and IgG isotypes **(Figure 2b)**, as has been previously reported for saliva and serum immunoglobulins ^26-28^.

**Figure 2.**
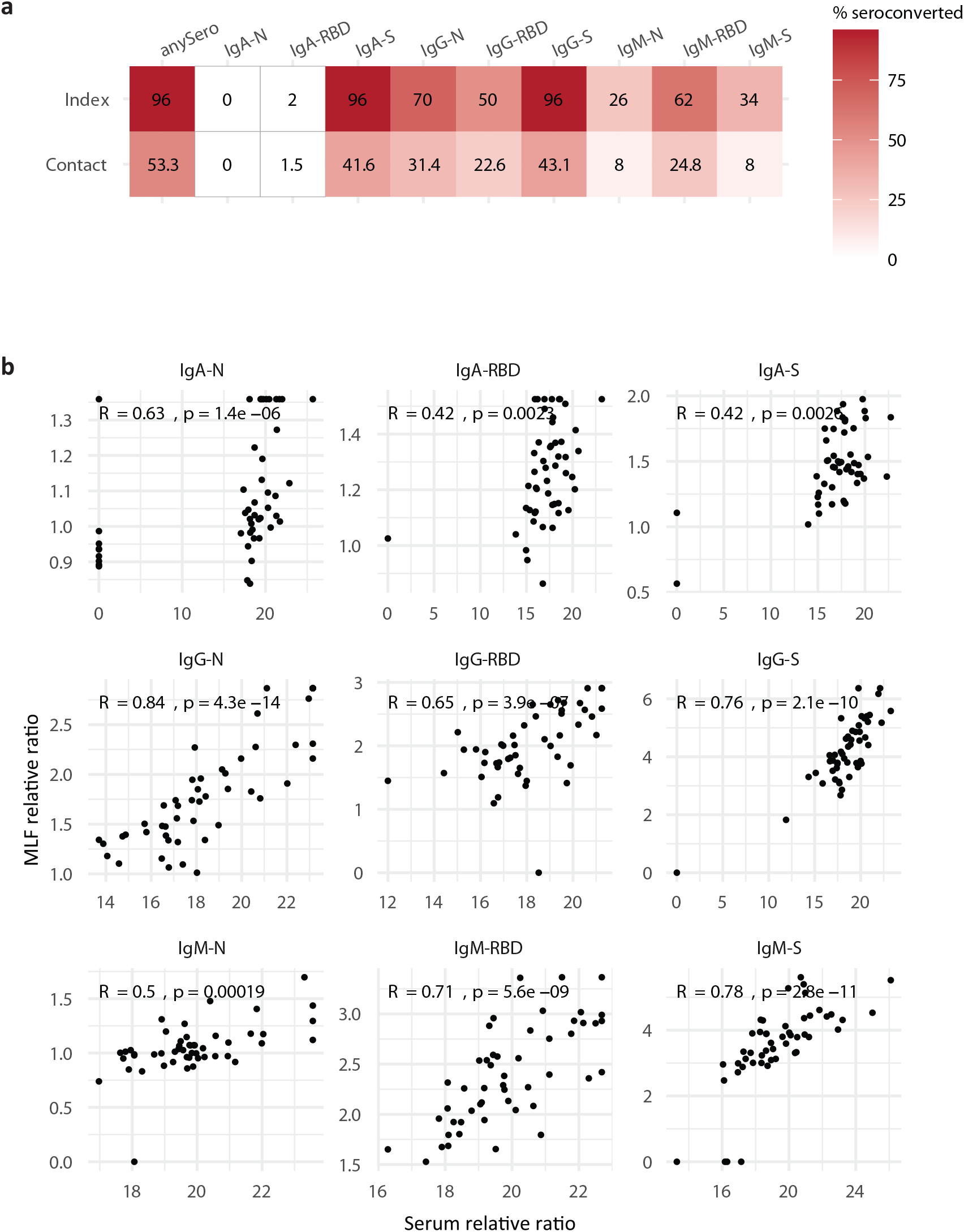
Serum antibody responses against SARS-CoV-2 in COVID-19 patients and household contacts. **a)** Seroconversion heatmap for index cases (N=50) and contacts (N=129) for each antigen and antibody isotype measurement, as well as seroconversion rate for any single antibody measurement (anySero). Seroconversion threshold was calculated as the mean of the controls plus 2∗standard deviation. **b)** Serum antibody responses against SARS-CoV-2 in COVID-19 in index cases correlated with mucosal antibody responses. IgM, IgG, and IgA antibody responses against SARS-CoV-2 spike protein (S), receptor binding domain (RBD) and nucleocapsid (N) were measured in sera or in mucosal lining fluid at the end of the study. Antibody levels are expressed as a ratio compared to the mean of pre-corona samples (relative ratio). Data are shown for index cases (N=50). Spearman correlations were calculated and p-values are reported directly in the figure.

### Mucosal antibody responses against SARS-CoV-2 in index cases

To examine the relationship between mucosal antibodies and virologic and clinical features, we focused on the index cases (n=50), as this group reported a date of symptom start. All index cases were included within two weeks of first experiencing symptoms **<u>(Figure S2a)</u>**, and all data of the index cases were collected between 0 and 40 days post symptom onset (PSO) **<u>(Figure S2b)</u>**, making the index cases a suitable cohort to examine these relationships during the acute phase of COVID-19 disease. The viral load at study enrolment was non-significantly lower when the individual had a longer period of symptoms before study start (**<u>Figure s3</u>**). We correlated MLF antibody measurements collected at study start with viral load and the days since symptom onset. A higher mucosal IgA antibody level at enrolment was correlated with a longer period of symptoms before study start, while viral load was negatively correlated with IgM levels specific for RBD and S antigens **(Figure 3a)**. This correlation pattern persisted after adjusting for age, sex, days since symptom onset for the viral load correlation, and viral load for the days since symptom onset correlation (data not shown).

**Figure 3.**
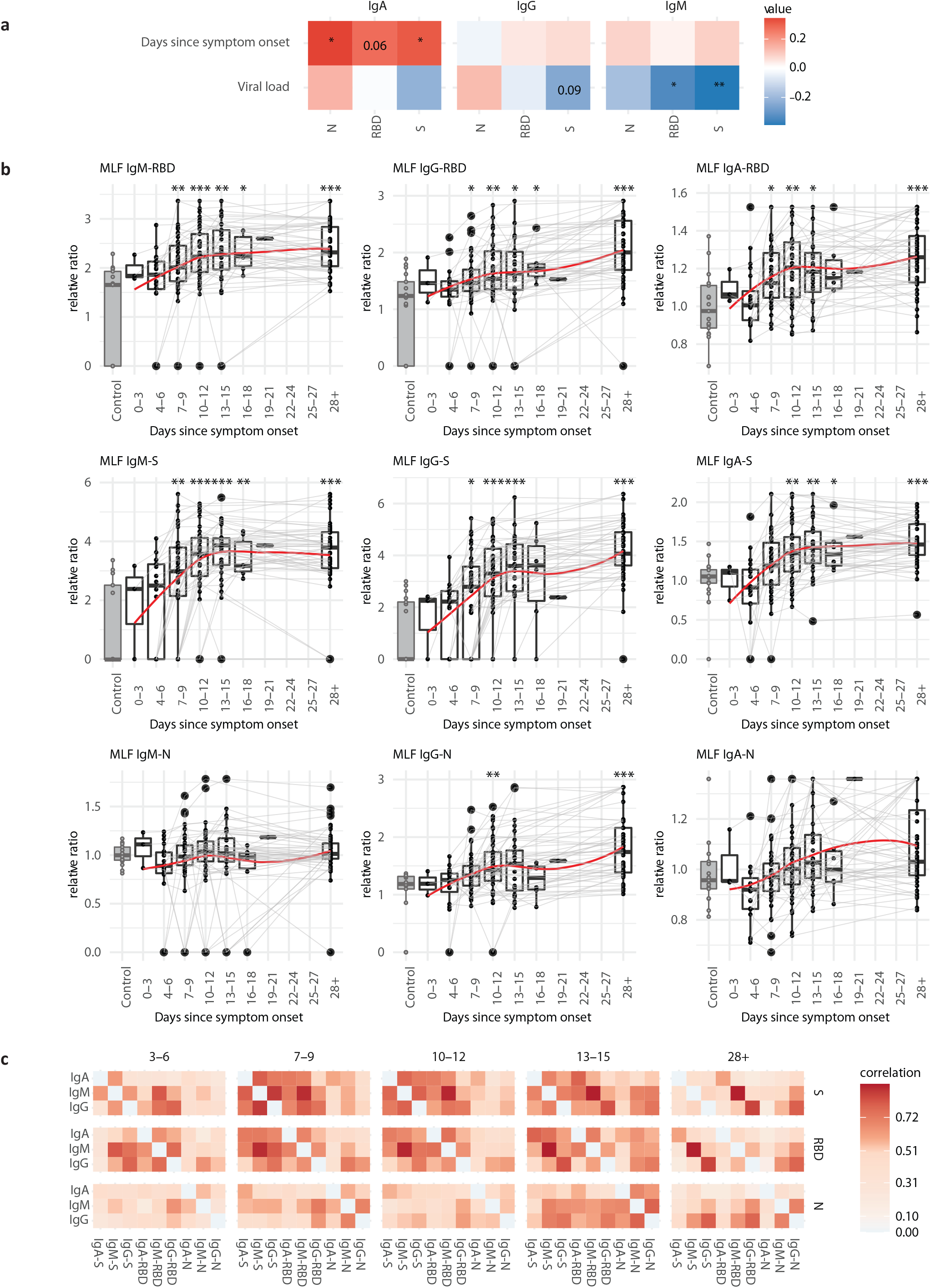
Mucosal antibody responses during SARS-CoV-2 infection in index cases (N=49). **a)** IgM, IgG and IgA antibody responses against SARS-CoV-2 spike (S), receptor binding domain (RBD) or nucleocapsid (N) collected in mucosal lining fluid at d0 were correlated with days since symptom onset (PSO) and viral load. Spearman correlations were calculated, ∗ p <0.05, ∗∗p<0.01, ∗∗∗ p<0.001. p-values of borderline significant tests are reported directly in the figure. **b)** Longitudinal mucosal antibody responses to S, RBD and N, plotted relative to the days PSO. Pre-SARS-CoV-2 controls are presented in the grey boxplots for comparison (n=17). Values within each timeframe were compared with the controls with the Wilcoxon-signed rank test. A non-parametric Loess curve is shown as red line to visualize the trend over time. Measurements from the same individual are connected with a grey line. **c)** Antibody measurements within the depicted timeframes were correlated with each other. Spearman correlation coefficient is presented.

To examine the kinetics of the mucosal antibody response, we measured the magnitude of antibody responses in the mucosal lining fluid of index cases at study day 0, 3, 6, and 28. Previous studies have shown that coronavirus-specific antibodies are significantly elevated in serum around two weeks PSO ^26,27,29^. We examined longitudinal mucosal antibody responses by binning samples into three-day timeframes relative to the day of symptom onset and plotting the values alongside controls. Mucosal IgM, IgA, and IgG antibody levels for S and RBD antigens were significantly elevated relative to controls between 7-9 days PSO and remained high up to 40 days PSO, while for N protein only IgG antibody responses were significantly higher than controls after 28 days PSO **(Figure 3b)**. In order to study the temporal changes in the relationship between the antibody-antigen types, we calculated correlations among antibody responses within each timeframe. While IgA, IgG and IgM isotypes were highly intercorrelated between S and RBD near the start of symptom onset, at the final measurement (28+ days POS) correlations between the IgA-S and IgA-RBD isotype were much weaker. Correlations of N-protein antibodies with S and RBD were overall weaker until at least two weeks PSO, at which point the IgG-N response correlated with the IgG-RBD and IgG-S antibody measurements **(Figure 3c)**.

Thus, SARS-CoV-2 infection induces a robust IgA, IgG, and IgM mucosal antibody response primarily against the S and RBD antigens as early as 7-9 days post symptom onset, while antibodies against N-protein show a different, slower induction and are overall restricted to IgG.

### Symptom progression in COVID-19 patients

Mild COVID-19 disease is highly variable in its presentation and is generally characterized as a SARS-CoV-2 infection that does not require hospitalization or oxygen support and is not accompanied by severe symptoms like pneumonia^4,8,30^. We examined the progression of 23 symptoms using a survey that all volunteers filled in daily throughout the follow-up. To analyse the clinical manifestation of mild COVID-19 disease over a period soon after symptom onset, we focused on the index cases. The most frequently reported symptoms across the entire study period were fatigue, rhinorrhea, coughing, headache, and anosmia **(<u>Figure S4a</u>)**. In order to compare different aspects of the clinical presentation, we grouped the symptoms into three categories: gastrointestinal symptoms (GS), systemic disease symptoms (SDS) and respiratory symptoms (RS). All index cases reported symptoms at the start of the study (**<u>Figure S4b</u>**). Fatigue was reported very frequently, and as such was not categorized **(<u>Figure S4a and b</u>)**. We examined whether the symptom duration varied between different clinical presentations, and calculated the duration for each index case for each symptom group. The duration of SDS was significantly shorter than RS, and by the end of the study period nearly all index cases had resolved their SDS, while a quarter still reported RS **(<u>Figure S4c</u>)**. To examine longitudinal changes in the magnitude of COVID-19 symptoms, we binned symptom notifications into three-day timeframes relative to the day of symptom onset, similar to the antibody analysis. While the number of SDS rapidly decreased from the time of symptom onset, RS appeared to stay high for at least two weeks post symptom onset **(Figure 4a)**. We investigated if patient characteristics were associated with changes in symptoms over time. We constructed a linear mixed-effects model per symptom group with the number of reported symptoms as the response and time since symptom onset, age, and sex as covariates. Such a linear model was a good fit for our longitudinal symptom data **(<u>Figure S5</u>)**. While time was significantly associated with decreases in symptoms, age was significantly associated with increased RS when correcting for the effect of time (p-value = 0.0165, **Figure 4b**). The sex had no effect on any symptom response group. To ascertain whether the induction of mucosal antibodies was associated with COVID-19 symptoms, the longitudinal mucosal antibody measures were added to the model in a univariate manner. This way, estimated changes in symptoms per unit increase of antibody signal represent the additional effect of antibodies while controlling for time, age, and sex. Increases in mucosal antibodies overall were associated with decreased SDS and RS. A significant association was only found for SDS, where high levels of IgM and IgG for S and all isotypes for RBD were related to a decrease in symptoms. The largest effect was seen in the relation between IgA-RBD and SDS **(Figure 4c)**.

**Figure 4.**
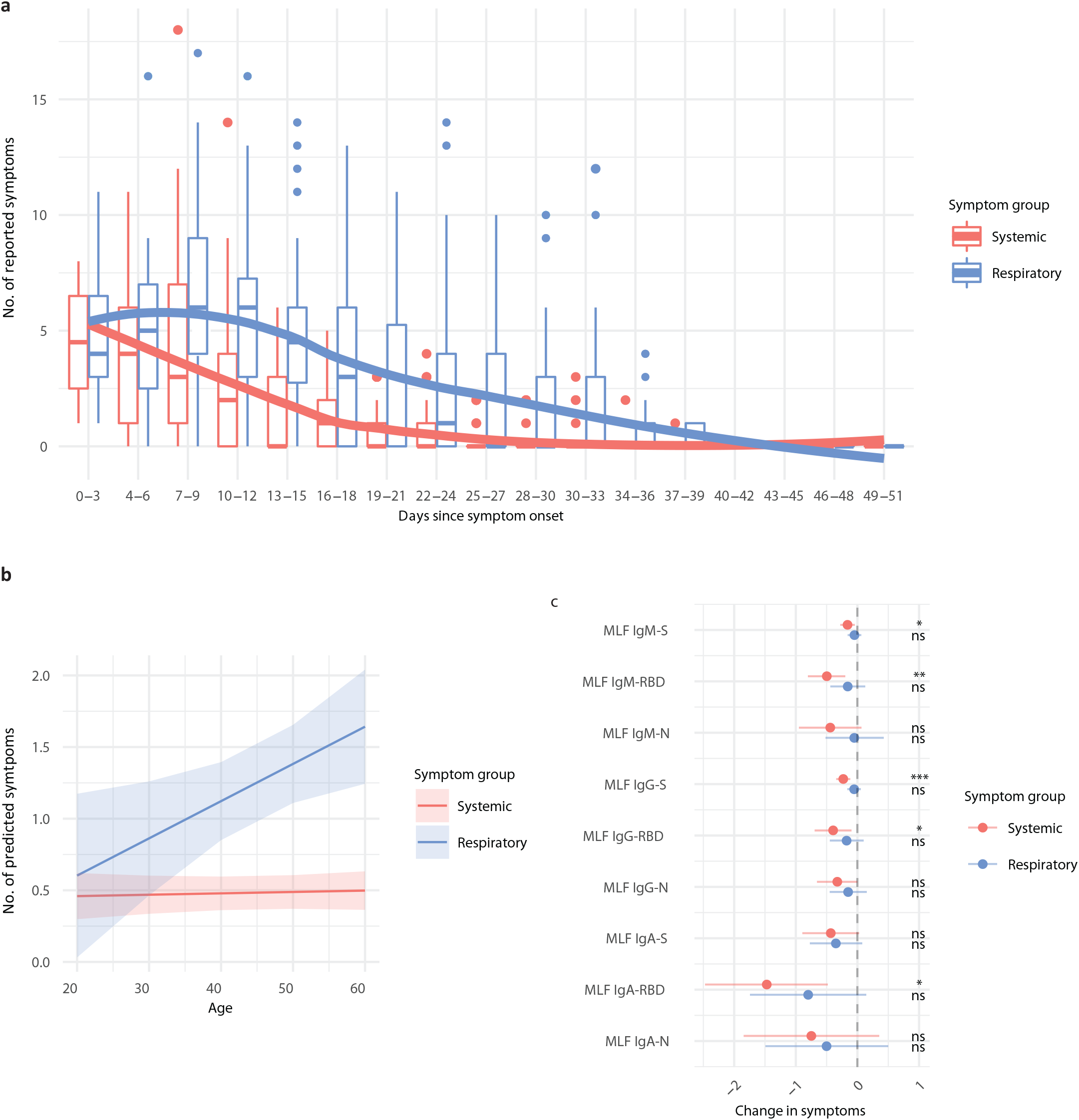
Influence of age and mucosal antibody levels on the progression of systemic and respiratory COVID-19 symptoms. **a)** The number of respiratory (RS) and systemic disease (SDS) symptoms were recorded for index cases (N = 49) for each day during the 28 day study period. Data are plotted relative to the time of symptom onset and values were binned into 3-day time frames. A non-parametric loess curve is shown as a red (SDS) or blue (RS) line in order to visualize the trend over time. **b)** A linear mixed-effect model (MEM) was fit to the data per symptom group. The response was specified as the number of symptoms on a given day, and explanatory fixed effects variables were: day since symptom onset, age, and sex. Time since symptom onset was also specified as a random slope, and Sample ID as a random intercept. A significant effect of age (p = 0.0165) was demonstrated for RS. Predicted symptom values from the model are plotted against index case age with 95% confidence interval bands. **c)** Longitudinal mucosal antibody measurements were added as fixed effect variables in a univariate fashion to the MEM from b). The predicted change in symptoms per unit increase of antibody signal is presented with 95% confidence intervals, and p-values for the association are plotted on the right (∗ p <0.05; ∗∗ p<0.01; ∗∗∗ p<0.001).

### SARS-CoV-2 infections in household contacts

To examine infections amongst household clusters, we classified household contacts into cases or non-cases. Cases were classified first by their PCR result on enrolment, followed by seroconversion for one of the antibody isotypes against S-protein and mucosal antibody responses against S-protein at the end of the follow-up. The PCR positive threshold was set at a Ct value <36, corresponding to a viral load of ≥10^3^ copies/ml. The seroconversion threshold was based on the mean + 2∗SD of the log2-transformed pre-SARS-CoV-2 control samples. To identify cases based on mucosal antibodies against S, we used a naive Bayes model that was trained by the mucosal antibody measurements of the PCR positive study participants (cases) and the pre-SARS-CoV-2 control samples (controls). Together, this approach identified 75 cases amongst 137 (54.7%) household contacts (**Figure 5a**), of whom 64% were PCR positive at study start, 87% were seropositive at the end of the follow-up, and 76% were identified as a case based on the MLF antibody levels at the end of follow-up (**<u>table S2</u>**). The majority of PCR positive cases also developed serum and MLF antibodies at the end of the study (88%). By including antibody measurements, 27 additional cases were identified (36% of cases).

**Figure 5.**
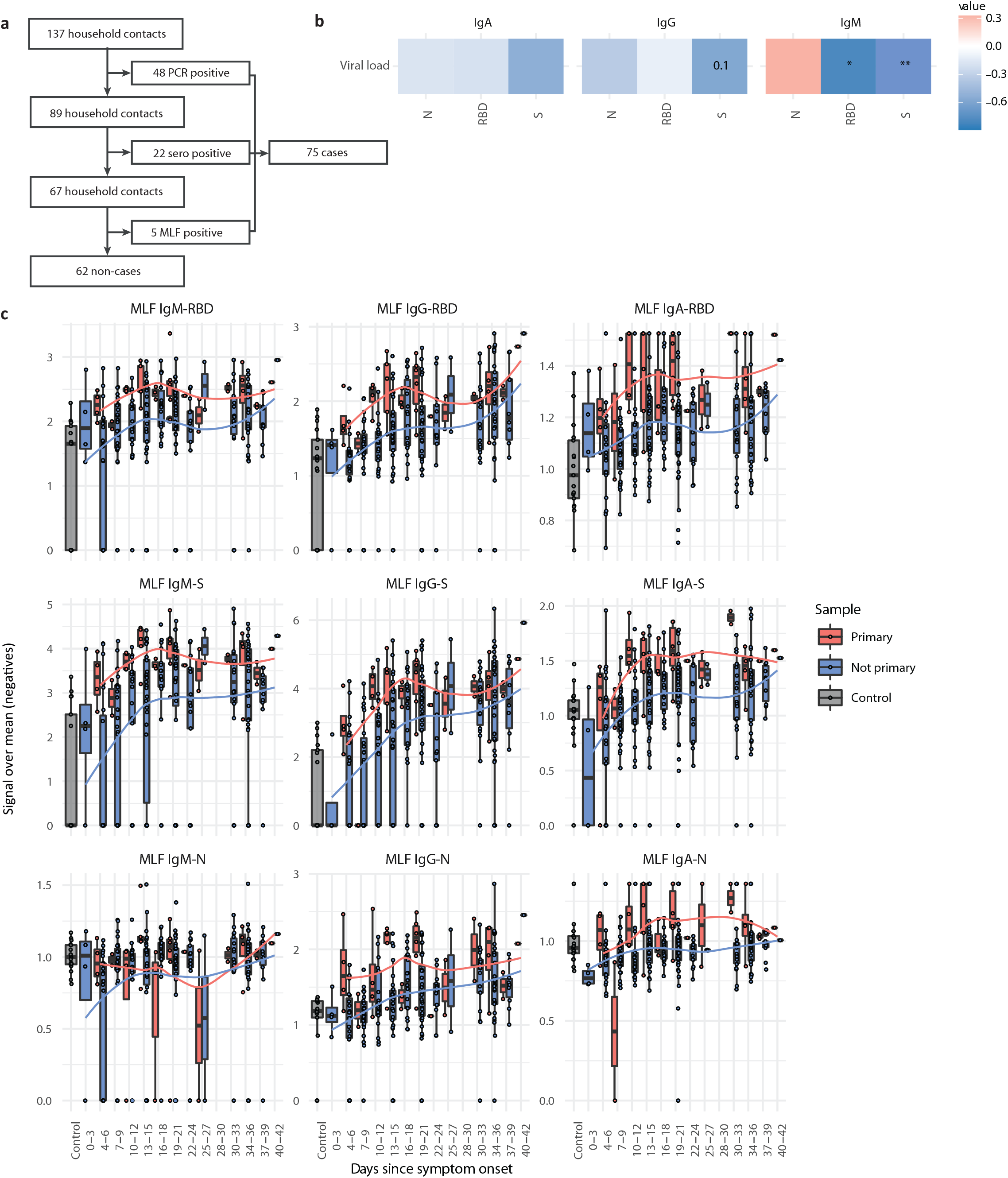
Identification of household contact cases and assessment of their mucosal SARS-CoV-2 antibody responses. **a)** Contact cases were classified into case or non-case first by their PCR result on enrolment, followed by seroconversion for either IgM, IgG or IgA against the S-protein, and mucosal antibody levels for either IgM, IgG or IgA against the S-protein at the end of the follow-up. The PCR positive threshold was set at a Ct value <36. The seroconversion threshold was based on the mean + 2∗SD of the log2-transformed pre-SARS-CoV-2 control samples. To identify cases based on mucosal antibodies against S, we used a naive Bayes model that was trained by the mucosal antibody measurements of the positive study participants (cases) and the pre-SARS-CoV-2 control samples (controls). **b)** Contacts were classified by their household as either the primary (N=9) or a non-primary (N=58) case, based on time of symptom onset. Mucosal antibody responses of the primary household contact cases collected at d0 were correlated with viral load. Spearman correlations were calculated, ∗ p <0.05, ∗∗ p<0.01, ∗∗∗ p<0.001. p-values of borderline significant tests are reported directly in the figure. **c)** IgM, IgG and IgA antibody responses against Spike protein (S), receptor binding domain (RBD) and nucleocapsid (N), measured in mucosal lining fluid. Data are plotted relative to the time of symptom onset of the index case in the respective household, and pre-SARS-CoV-2 control samples are presented for comparison. A non-parametric loess curve is shown as a red (primary cases) and a blue (non-primary cases) line to visualize the trend over time.

All participants were asked if they were the first with COVID-19 related symptoms (primary case) in the household (parents were asked to answer for young children). This classification was solely based on symptoms, as only healthcare workers were eligible for PCR testing at the time of this study. Among the contacts included in the study, nine were reported to be the primary case based on the timing of symptom onset. When we correlated their mucosal antibody responses to viral load, a similar negative correlation of IgM levels with viral load was observed for the S and RBD antigens as in the index cases (**Figure 5b**). We hypothesized that the primary cases within the household contacts would have been symptomatic for a longer period than the non-primary contacts. To see if this different stage of infection would be reflected in a difference in mucosal antibody levels, we plotted their mucosal antibody values in a similar manner as the index cases, using the day of symptom onset of the index case in the respective household as we did not have this data for the contacts. We found that primary cases, compared to secondary cases, had earlier and overall higher antibody responses for all antibody isotypes of S and RBD antigens (**Figure 5b**). Similar to the index cases, IgM and IgA antibody responses against the N-protein were relatively weak and were in range of the pre-SARS-CoV-2 control samples.

The role of children in transmission of SARS-CoV-2 has been the subject of significant discussion. We therefore examined the likelihood that children <12y become infected. We found that 16 of the 33 (48.5%) of the children <12y were infected, which was not significantly different compared to the 59/104 (53.8%) in the >12y household contacts (p-value:0.78, **Figure 6a**). In the contacts above 50 years, the proportion of infection was the highest (19/ 31, 61%). Most of these infected contacts above 50 years were the partner of the index case (12/19, 63%), while all infected participants <12y were children of the index case. Of note, 29% of all infected household members did not have any symptoms at study start, and 19% of all infeceted contacts remained asymptomatic throughout the entire follow-up. When examining the distribution of asymptomatic cases among the four age groups, we found that 7 out of the 16 (44%) cases <12y were asymptomatic at study start, compared to 11 of the 59 (20%) cases above 12y (p-value:0.10, **Figure 6b**). When we examined the overall number of symptoms that was reported throughout the follow-up, we noticed a strong and significant positive correlation between number of symptoms and age (**Figure 6c**). Despite the lower number of symptoms or absence of symptoms in child cases, no differences were found in viral load compared to the older age categories, nor were the serum antibody levels at the end of the follow-up different between the age-groups (**Figure s6a and b**).

**Figure 6.**
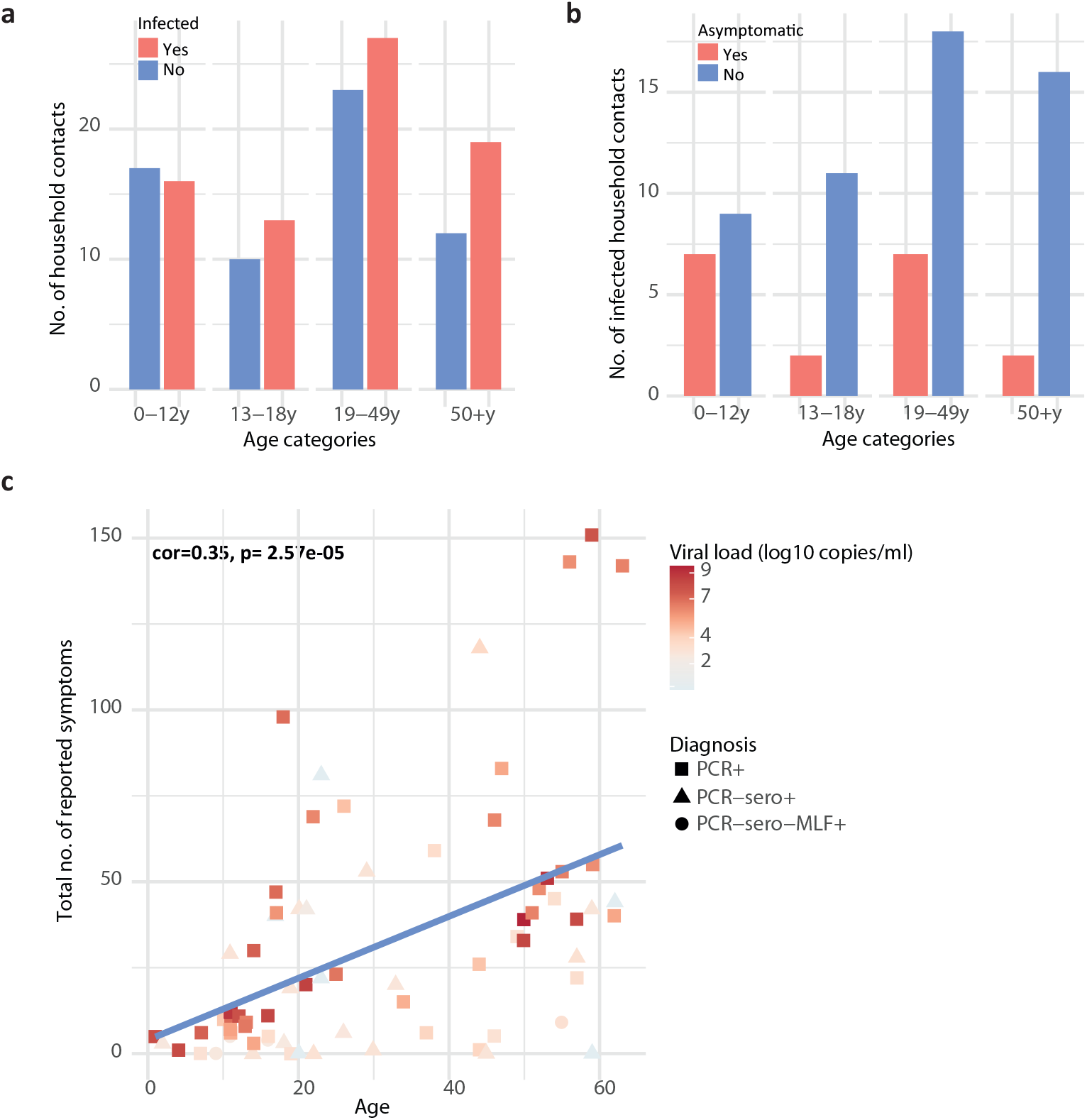
SARS-CoV-2 infection, viral load and number of symptoms of contact cases, split into four age groups. **a)** Proportion of infected household contacts per age group. Sixteen out of 33 primary school-age children (1-12y) were infected (48%), 13 out of 23 children aged 13-18y (56%), 27 out of 50 adults aged 19-49 years (54%) and 19 out of 31 contacts older than 50 years (61%). **b)** Proportion of asymptomatic disease among household cases (N=75) at study enrolment within each age category. Seven of the children below 12 years were asymptomatic (44%), two of the children up to 18 years (15%), seven of the adults up to 49 years (28%) and two of the contacts older than 50 years (11%). **c)** The correlation between the total number of reported symptoms and the age of the infected household contacts. Pearson’s correlation and significance is depicted in the figure. The shape of the datapoints represents the method by which a contact was classified as a case and the color of the datapoints shows the viral load at study start.

## DISCUSSION

In this study, we examined the mucosal antibody responses following infection with SARS-CoV-2 and the development of mild COVID-19 disease in 50 adults and their household contacts. This study demonstrates the unique value of using mucosal lining fluid to assess various aspects of SARS-CoV-2 infection, ranging from pathophysiology to epidemiology.

Overall, we observed a significant increase in antibody levels measured in the mucosal lining fluid of the index cases, with a timing that is similar to what has previously been described for serum and saliva ^6,26,27^. Mucosal IgG and IgM antibody responses measured against the three viral antigens correlated strongly with serum antibody levels, but were weaker for IgA, measured at 28-40 days PSO. This has previously been described for saliva^26,27,31^. A key finding of this study is that the mucosal IgA response against RBD showed the strongest relation to resolution of systemic disease symptoms. This phenomenon has also been addressed by Butler *et al*.^32^ in a smaller cohort (n=20). They found that strong mucosal IgA responses against SARS-CoV-2 was related to neutralizing activity and that elevated levels of mucosal neutralization were associated with mild and moderate symptoms. This neutralizing activity of IgA has also been described in a bigger study that mainly focused on serum^33^. Another study in one family showed spiking salivary IgA levels at the time of symptom resolution, while serum antibody levels remained low.^3^ Our larger study, including for the first time the dynamic measurement of the lining fluid of the upper respiratory tract, clearly shows the relation of mucosal antibodies with a decrease in symptom burden. Nevertheless, we did not measure neutralizing activity, and found that other mucosal responses against S and RBD also contributed to a reduction of systemic disease symptoms. Another significant finding was the negative correlation between viral load and mucosal IgM against RBD and S, measured at study start, suggestive of a neutralizing role of mucosal IgM, although other possible explanations, like pre-existing T-cell immunity cannot be excluded^34,35^.

Thus, a strong mucosal antibody response might play an important role in preventing severe disease, by reducing the systemic symptoms and reducing viral load. Patients with severe COVID-19 are sometimes treated with convalescent plasma from patients who have recovered from COVID-19^36,37^. Given their multimeric nature, IgM and IgA antibodies specific for RBD are likely to neutralize SARS-CoV-2 particles and strong neutralizing capacity has been described previously for both isotypes^33,38^. Therefore we propose that these antibodies should be included in the quality evaluation of convalescent plasma. Moreover, monitoring of SARS-CoV-2-specific antibody concentration in MLF in patients receiving convalescent plasma may yield important insights into which doses are required to reach sufficient concentrations at the primary site of infection. Intranasal administration of SARS-CoV-2 specific IgM or IgA in patients recently diagnosed with COVID-19 may provide an additional treatment option.

Because we measured the mucosal response of three antibodies against three different antigens, we were able to describe correlations between these antigen/isotype combinations. We found that there was a coherent antibody production against S and RBD early in the infection. Correlations between S and RBD with the N antigen were significantly less pronounced, and only emerged later in the infection, mostly for IgG. This indicates that the antibody response to the nucleocapsid antigen follows a different track after infection, than S and RBD, which was also seen in previous studies^26,27,31^. This variation in the antibody response composition could be explained by the cross-reactivity with other seasonal coronaviruses. Previous exposure with these coronaviruses could elicit a booster response instead of a primary immune response during SARS-CoV-2 exposure, explaining the presence of IgG and IgA and absence of IgM, which has the strongest response after primary exposure. Next to this, the release of N-proteins requires the lysis of virally infected cells, as it encapsulates the viral RNA. This could explain the difference in timing and the generally low mucosal antibody responses against N, as all our participants developed mild disease.

The ability to measure the mucosal antibody response over a longer period of time in an easy, non-invasive manner allowed us to identify cases among the household members that were not picked up by PCR-testing at a single timepoint only. When we performed the study, we were unable to collect additional samples for viral PCR due to shortages in swabs and transport medium, limiting our ability to fully study the dynamic interactions between viral infection and antibody responses, and calculate the sensitivity of the antibody measurements in diagnosing infection in comparison to PCR. We observed a very high proportion of cases among the household contacts (54.7%), which is much higher than previously reported (11-37%) ^39-41^. This can partly be explained by the fact that not all the index cases were the primary case, due to the restricted PCR testing on specific risk-groups at the time the study was performed. By using the households’ primary case reporting, the secondary attack rate (SAR) was lowered to 49.6%. The fact that we still find a higher SAR than described in previous studies may be explained by the fact that almost all household transmission studies to date identified cases based on PCR positivity only, and thus underestimate the true number of cases within a cluster. PCR testing is prone to false negative results due to it being a one-time measurement, its dependency on the time of sampling in relation to the infection, and the chance of errors during sample collection. By combining PCR, serology and mucosal data, we were able to identify an additional 27 cases, increasing the total number of contact cases by a third. Although we used the mucosal antibody levels as an exploratory case-identification tool and used PCR and serology for most of the case identifications, the majority of serum and PCR positive contacts also had mounted a mucosal antibody response towards the end of the follow-up (**<u>Table S2</u>**). These results suggest that mucosal lining fluid could be used as a non-invasive way to measure disease exposure in a community setting, and as a tool to identify the primary case in a household.

Children were initially thought to be play less of a role in SARS-CoV-2 infection and transmission, because of a lower expression of ACE2 and the high frequency of asymptomatic infections compared to adults ^42-44^. This notion has been debated in research^11,45-48^, and is critical for making decisions related to epidemic control measurements, like school-closure. In our study, we found that children <12 years had a similar probability to become infected as adults. Almost half of the child cases in our study were asymptomatic, their viral load at study start was similar to that of adults, and other studies have shown that 40-80% of transmission events take place before symptom onset ^49-51^. A study looking at the viral shedding of asymptomatic, presymptomatic and symptomatic children did not find a difference in viral shedding between these groups^52^. Together, these findings imply that children may play a larger role in transmission of SARS-CoV-2 than is currently thought. A prospective cohort study in which MLF samples are collected irrespective of symptom presentation would provide more insight in whether (asymptomatic) infections in children are an important driver of household transmission or not. Since collection of MLF for antibody analysis is rapid, non-invasive and can be done by study participants in the absence of health care professionals, such studies including repetitive sampling are feasible in this age group.

Our study has several limitations. First, the starting point for our study was the inclusion of healthcare workers, most of whom were female, and not entirely representative for the larger population. It should be noted that this study was performed during the first wave in March-April of 2020, when all schools in the Netherlands were closed (**Figure 1b**), minimizing contacts between children and thus making it hard to study the role of child-child contacts in the transmission of SARS-CoV-2. Lastly, additional mucosal lining fluid measurements would have enabled more advanced modelling of the mucosal antibody trajectories, especially in the household contacts.

This study provides an example of the unique possibilities of studying mucosal antibody trajectories. It provides essential new insights into the mucosal antibody kinetics after a SARS-CoV-2 infection, and uncovers novel relations with viral load and symptom resolution. Furthermore, the study design and analysis strategy presented here can be used as a blueprint for follow-up investigations not only for COVID-19, but also for other infectious diseases. The ability to collect repetitive mucosal antibody samples in a non-invasive manner removes an important obstacle for use in age groups that are normally difficult to sample, such as children.

## ONLINE METHODS

### Recruitment

This observational prospective cohort study was conducted among COVID-19 cases with a laboratory confirmed infection, as well as their household members that remained in home quarantine at the same address. The study was conducted in accordance with the provisions of the Declaration of Helsinki (1996) and the International Conference on Harmonization Guidelines for Good Clinical Practice. The study was approved by the local medical research ethics committee and is registered with ClinicalTrials.gov (NCT04590352; ethical committee reference NL73418.091.20). All index cases in this study were healthcare workers (HCW) from three hospitals (Radboudumc, CWZ and Rijnstate) in the provinces of Gelderland in the Netherlands, with a confirmed SARS-CoV-2 infection. Study participants were included from the 26^th^ of March 2020 until the 15^th^ of April, when the inclusion number of 50 households was reached. Participants were introduced to the study through the occupational health and safety services (OHS) of the participating hospitals. HCWs were included if they had a positive Polymerase Chain Reaction test (PCR-test) for the SARS-CoV-2 virus, tested and judged by the OHS of their hospital, with a positive indication for home isolation, and had at least two household members willing to participate.

### Study design

Before the first home visit, all index cases of the family had a telephone interview, where they were asked about their first day of symptom onset, whether they were in isolation from the rest of the household, whether physical contact was restricted with other household members, whether they were still symptomatic, and whether they thought they were the primary case in the household. Households were visited within 1-2 days of a positive PCR in the index case. Following informed consent, nasopharyngeal and oropharyngeal swabs were taken for viral PCR, as per diagnostic guidelines^53^. A nasal mucosal lining fluid (MLF) sample was obtained from all participants by the use of the Nasosorption™ FX·i nasal sampling device (Hunt Developments, UK). A synthetic absorptive matrix (SAM) strip was gently inserted into the nostril of the participant and placed along the surface of the inferior turbinate. The index finger was lightly pressed against the side of the nostril to keep the SAM strip in place and to allow MLF absorption for 60 seconds, after which the SAM strip was placed back in the protective plastic tube. Participants were instructed on how to self-sample MLF at home. Finally, participants were asked about their symptoms of that day.

Participants were followed up for approximately 28 days, starting on the day of the first home visit (day 0) and ending on the last home visit (day 28-33). This range in the last visit was due to logistical difficulties during the summer holidays; 14 index cases were visited on their day 29, three on day 30, five on day 31, four on day 32 and one on day 33. All particpants registered their symptoms for 28 days. During follow-up, clinical symptoms were registered three times daily and MLF was collected at three different study days via self-sampling. For the index case, MLF was collected on day 0, 3, 6 and 28-33 and for the household contacts this was on day 0, 7, 14 and 28-33 (**Figure 1c**). Self-sampled MLF samples were stored in biosafety bags in the participants’ own freezer (temperature around −20 °C).

At the final home visit, MLF samples were picked up and transported to the Radboudumc on dry ice, where it was stored at −80°C until further testing. For antibody analysis, Nasosorption™ FX·i nasal sampling devices were thawed on ice, after which the synthetic adsorptive matrix (SAM) was removed using sterile forceps. The SAM was placed in a spin-X filter Eppendorf tube with 300uL of elution buffer (PBS/1% BSA) for a minimum of 10 minutes, followed by centrifugation at 16000x*g* for 10 minutes at 4°C. To prevent unspecific binding, the spin-X filter columns were pre-incubated with the blocking buffer for 30 minutes.The filter cups were then removed from the Eppendorf tubes using sterile forceps. To inactivate live SARS-CoV-2, the eluate was incubated for a minimum of 45 minutes at 56 °C, spun down, aliquotted and stored at −20°C for further testing.

Finger-prick blood (∼0.3 ml) was collected from all participants consenting to the finger-prick at day 28 by the use of a sterile disposable lancet device (BD Microtainer Lancet) and a sterile capillary tube. Blood samples were kept at room temperature until processing at the Radboudumc laboratory site, after which serum was stored at −20°C until further testing.

All collected symptom diaries were digitalized into Castor EDC, clinical trial software for electronic data capture and clinical data management.

### Sample analysis

#### Detection of SARS-CoV-2

Presence or absence of SARS-CoV-2 and viral copy number per ml was determined on the combined nasopharyngeal and oropharyngeal swab, using a PCR protocol that was developed at the Radboudumc Medical Microbiology Laboratory. Nasopharyngeal and oropharyngeal swabs were collected in GLY medium and stored at −80°C untill processing. Samples were thawed, vortexed and 500 ul of sample was lysed in 450 ul MagNAPure Lysis/binding buffer (Roche). An ivRNA internal extraction control was added and samples were extracted on the automated MagNAPure LC2.0 system using the MagNAPure LC Total Nucleic Acid isolation kit - High Performance (Roche). Samples were eluted in 50 ul of which 5 ul was used in the RT-qPCR using the Luna Universal Probe One-Step RT-qPCR kit (NEB) with 400 nM E-gene primers (FW: 5’-acaggtacgttaatagttaatagcgt-3’ RV: 5’-atattgcagcagtacgcacaca-3’) and 200 nM E-gene probe (5’-FAM-ACACTAGCCATCCTTACTGCGCTTCG-BHQ1-3’ (Biolegio)) on a CFX96 Real-Time PCR Detection System (BioRad). Transcript quantities were calculated using a 10-fold dilution series of E-gene ivRNA. The extraction efficiency was checked in a separate RT-qPCR using the Luna Universal Probe One-Step RT-qPCR kit (NEB) with primers targeting the ivRNA that was added prior to extraction.

#### Antibody measurements

For antibody analysis, a fluorescent-bead-based multiplex immunoassay (MIA) was developed. The stabilized pre-fusion conformation of the ectodomain of the Spike protein (amino acids 1 – 1,213) fused with the trimerization motif GCN4 (S-protein) and the receptor binding domain of the S-protein (RBD) as previously described by Wang C. et al.^54^, and the Nucleocapsid-His recombinant Protein (N) (40588-V08B, Sino Biologicals), were each coupled to beads or microspheres with distinct fluorescence excitation and emission spectra, essentially as described in the paper by den Hartog *et al*.^55^

A total of six reference serum samples were selected from PCR confirmed COVID-19 patients with varying immunoglobulin G (IgG) concentrations, and pooled to create standard curves for IgG, IgA and IgM. Next to this, four different samples from the same cohort were used as quality control samples. As negative control samples, we used historical serum (n=32) and MLF (n=17) samples collected prior to the SARS-CoV-2 pandemic.

MLF samples were diluted 1:5 in assay buffer (PBS/1% BSA/0.05% tween-20) and serum samples were diluted 1:500 in assay buffer, incubated with antigen-coated microspheres for 30 minutes at room temperature while shaking at 450 rpm. Following incubation, the microspheres were washed two times with PBS, incubated with phycoerythrin-conjugated goat anti-human, IgG (Jackson Immunoresearch, 109-116-170), IgM (Southern Biotech, 2022-09) and IgA (Southern Biotech, 2052-09) for 20 minutes and washed twice. Data were acquired on the Luminex FlexMap3D System. Validation of the detection antibodies was obtained from a recent publication using the same antibodies and the same assay^55^, and specificity was checked using rabbit anti-SARS SIA-ST serum.

S- and N-coupled microspheres were combined to measure antibodies directed against multiple antigens (or epitopes) in one single sample. Since antibodies against the S-protein and RBD may compete for the same epitopes, antibody binding to RBD was measured separately. Using different conjugates, IgG, IgA, and IgM-specific antibodies concentrations were measured in MLF and serum. MFI was converted to arbitrary units (AU/ml) by interpolation from a log-5PL-parameter logistic standard curve and log-log axis transformation, using Bioplex Manager 6.2 (Bio-Rad Laboratories) software and exported to R-studio. Negative control samples (MLF and serum) were used to filter out background signal in the antibody measures. The MLF samples originated from the KIRA-study performed at the Radboudumc, in which healthy healthcare workers are vaccinated against pertussis as per routine care, and gave consent to the use of the MLF samples for other research. The serum samples originated from the Radboudumc Biobank, that allows the use of serum samples for research as long as privacy of the donors is guaranteed. The standard dilution range plus four quality control samples were added to each plate.

### Symptom categorization

To analyse the relation between symptom clearance in index cases and the mucosal antibody response, we categorized our set of symptoms into three categories, based on their clinical presentation. This resulted in a set of 23 symptoms, which were categorized into three categories, i.e. respiratory symptoms (RS) systemic disease symptoms (SDS), and gastrointestinal symptoms (GS). RS includes chest pain, sneezing, nose bleeding, pain when breathing, coughing with mucus, dyspnoea, sore throat, loss or change of taste/smell (dysnosmia), coughing, and rhinorrhoea. SDS includes dizziness, headache, fever, temperature, chills, joint pain, muscle pain, swollen lymph nodes and low appetite. GS includes vomiting, diarrhoea, and nausea. Because fatigue was reported in almost all cases, we did not categorize it into one of the symptom categories.

### Case definition and cluster identification

For analysis of SARS-CoV-2 exposure within households, we categorized the household contacts into cases and controls. Cases were defined as being either PCR positive at study start and/or seropositive for IgA, IgG or IgM against S at the end of follow-up. PCR positivity was set on a Ct value<36, which corresponds to a viral load of at least 10^3^ copies/ml extracted sample. The seroconversion threshold was based on the mean + 2∗SD of the historic negative control samples, which were collected before SARS-CoV-2 was introduced in the Netherlands. Additionally, we used a Naïve Bayesian probabilistic model (R-package “naivebayes”, with prior probabilities derived as the class proportions for the training set, and Laplace smoothing set at 1) to identify cases within the PCR/seronegative contacts by the anti-S MLF antibody response on day 28. Mucosal antibody levels from PCR positive cases were used as positive controls; mucosal antibody levels from the historic cohort samples as negative controls. These values were used to train the probabilistic model, after which the model was applied to the PCR/seronegative contacts.

### Statistical analyses

Analysis of Luminex data was performed with Bio-Plex 200 in combination with Bio-Plex manager software (Bio-Rad Laboratories, Hercules, CA). Demographical data was exported from Castor EDC, and double checked with the paper records by two members of the research team. All statistical analyses were performed using the Rstudio environment, with libraries ‘stats’ (hypothesis tests and correlations), “naivebayes” for the naïve Bayes classification, “lme4” ^56^, “lmerTest” ^57^ for mixed-effects modelling and associated p-values, and “survival” ^58^ for Kaplan-Meier survival analysis. The libraries “survminer” and “ggplot2” were used for visualization. Changes in serum or mucosal antibodies compared to negative controls were tested using a two-tailed paired Wilcoxon signed rank test, and then corrected for multiple testing with the Benjamini-Hochberg method ^59^. Statistical parameters including the sample sizes, measures of distribution, and p-value thresholds for significance are reported directly in the figures and figure legends. In order to determine if a sample was seropositive for a given combination of antigen and antibody isotype, a cut-off value (mean + 2 standard deviations) was calculated from the negative control samples. Samples above this threshold were classified as seropositive for that antigen and isotype combination. Samples that were seropositive for any of the antibodies tested were classified as such (“anySero”, Figure 2A). Where correlations are presented, the spearman correlation coefficient and associated p-value were calculated. The time at which a subject became symptom-free was calculated as the last day that individual showed symptoms for that category, plus one day. The probability of becoming symptom-free was estimated using Kaplan-Meier’s method, and the hypothesis testing was performed using log-rank test. In order to estimate the effect of patient characteristics and antibodies on symptoms over time, we constructed a mixed-effects model. For each subject and for each timepoint, we added together the number of complaints per symptom category. We specified a mixed-effects model per symptom category with symptoms as the response and time since symptom onset (POS), age, and sex, as explanatory variables. We also added POS and Sample_ID as random effects. The formula for the model (in R notation):

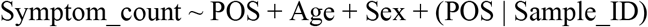

In order to determine the effect of antibodies on the symptom response, the model above was updated in a univariate fashion with each antibody measurement as a covariate. The formula of the updated model:

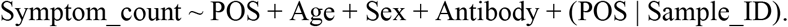

Estimates for the covariates, as well as 95% confidence intervals and p-values (Satterthwaite’s approximation to degrees of freedom) were extracted and plotted.

## Data Availability

The data and R-code that support the findings of this study are available from the corresponding author upon reasonable request.

## FUNDING

This study was funded by the Laboratory of Medical Immunology, Radboud university medical center.

## CONTRIBUTORS

Conceptualization: D.A.D and M.I.J.; investigation: J.F., D.A.D., R.P., J.R., D.T., and M.B.; lab processing: J.F., R.P., D.A.D., K.L., E.S., and C.E.G.-J.; formal analysis: J.F. and J.G.; writing— original draft preparation: J.F., and J.G.; writing—review and editing: J.G, D.A.D., T.B., M.A.H., M.I.J., and R.P.; resources: F.J.K, B.-J.B., N.G.-B, C.D., and M.N.-F.; and supervision: D.A.D.

## DECLARATION OF INTERESTS

There are no competing interests to be declared.

## ACKNOWLEDGEMENTS

We would like to thank all MuCo study participants for their willingness to participate and their support for the study. Additionally, we thank M. Boonstra, M. Blok, D. van der Giessen, E. Lenssen, E. Reuvers, M. Roek and E. Wijnhoven for assisting with performing the first home visit, H. Lyoo from Utrecht university for experimental assistance, J. Heijnen, M. Dautzenberg, and A. Voss for recruiting participants from the occupational health and safety officers at Rijnstate and CWZ hospitals, and the respective medical microbiology departments involved in PCR testing of hospital employees.

## FIGURE LEGENDS

**Figure S1. Serum antibody responses against Sars-CoV-2 in COVID-19 patients and household contacts**. IgM, IgG, and IgA serum antibody responses against Sars-CoV-2 spike protein (S), receptor binding domain (RBD), or nucleocapsid (N). Data are shown for pre-SARS-Cov-2 control samples (N=32), index cases (N=50), or household contacts (N=129). Antibody levels are expressed as a ratio compared to the mean of the controls (relative ratio) and values of indexes and contacts were compared to those of controls using a Wilcoxon signed rank test, ∗ p <0.05; ∗∗ p<0.01; ∗∗∗ p<0.001, and a threshold for seroconversion (dotted line) was calculated based on the mean + 2∗sd of the control samples.

**Figure S2. Timing of COVID-19 patient recruitment and sample measurements relative to symptoms onset. a)** The graph shows the cumulative percentage of households (N=50) included at a given time post symptom onset. **b)** Overview of the measurements made for the index cases relative to the reported first day of symptom onset. Each line represents one participant.

**Figure S3. SARS-CoV-2 viral load at different days post symptom onset**. Index cases (N=49) were asked when their symptoms started, and viral load was measured at study start. Individuals are binned together based on their reported symptom onset, relative to the study start. The y-axis shows the average viral load in log10 copies/ml extracted sample.

**Figure S4. Clinical presentation of mild COVID-19 disease. a)** Index cases (N = 49) completed a daily symptom survey covering 23 symptoms for 28 days during the study period. The number of index cases that reported a given symptom at any time during the study period is presented. Symptoms were categorized into either respiratory symptoms (RS), systemic disease symptoms (SDS), or gastrointestinal symptoms (GS). Fatigue was not categorized. **b)** Cumulative symptoms per symptom group for each study day are plotted for each index case. Data are represented since the time of symptom onset. A window depicts the study period for a given index case. **c)** Symptom durations were calculated for RS and SDS, and compared using a Kaplan Meier analysis. The probability of becoming symptom-free at a given time post-symptom onset is depicted. The log-rank p-value represents the statistical difference between resolution of SDS and RS.

**Figure S5. Mixed-effects modelling of longitudinal symptom data in COVID-19 patients**. The number of respiratory (RS) and systemic disease (SDS) symptoms were determined for index cases (N = 49) for each day during the 28 day study period. Data are represented relative to the time of symptom onset. A linear mixed-effect model was fit to the data per symptom group. The response was specified as the number of symptoms on a given day, and explanatory fixed effects variables were: day since symptom onset, age, and sex. Time since symptom onset was also specified as a random slope, and Sample ID as a random intercept. Datapoints are plotted per day and joined by a faded line, predicted values from the mixed-effects model are plotted on top as solid straight lines.

**Figure S6. Viral load and anti-Spike serum antibody concentrations in household contact cases of different age categories. a)** Viral load at study enrolment, depicted for the PCR positive household cases (n=46). The y-axis shows the average viral load in log10 copies/ml extracted sample of the contact cases, the x-axis shows the four different age categories. **b)** Serum anti-Spike protein levels at the end of the study, for IgA, IgM and IgG, in each age category. Serum antibody levels are depicted as the log10 antibody concentration in AU/ml. Only infected household members were selected (n=75).

## Notes

### Competing Interest Statement

The authors have declared no competing interest.

### Clinical Trial

NCT04590352

### Author Declarations

The study protocol was approved by the Arnhem-Nijmegen Medical Ethical Committee, is registered at clinicaltrials.gov (NCT04590352; ethical committee reference NL73418.091.20), and was performed in accordance with the declaration of Helsinki. Written informed consent was obtained from volunteers before any research procedure was initiated.

