## Supplemental Table 1 and 2 for "Elevated mucosal antibody responses against SARS-CoV-2 are correlated with lower viral load and faster decrease in systemic COVID-19 symptoms"

| Variable | Index cases (n=50) | Household contacts (n=137) |
|---|---|---|
| **Age** median [IQR] | 46 [37-54] | 21 [13-46] |
| **Age groups** n(%) | | |
| *0-12 years* | - | 33 (24) |
| *13-18 years* | - | 23 (17) |
| *19-49 years* | 31 (62) | 50 (36) |
| *50+ years* | 19 (28) | 31 (23) |
| **Sex Female** n(%) | 38 (76) | 54 (39) |
| **Diagnosis** n(%) | | |
| *PCR+* | 46 (92) | 48 (35) |
| *PCR- Sero+* | 3 (6) | 22 (15) |
| *PCR- Sero- MLF+* | 1 (2) | 5 (4) |

**Table S1 | General characteristics of study participants.**

| | Index cases (n=50) | Household cases (n=75) |
|---|---|---|
| **PCR +** | **46 (92)** | **48 (64)** |
| *PCR+ sero+ MLF+* | *43 (86)* | *42 (56)* |
| *PCR+ sero+ MLF-* | *2 (4)* | *1 (1)* |
| *PCR+ sero- MLF+* | *0* | *1 (1)* |
| *PCR+ sero- MLF-* | *1 (2)* | *4 (5)* |
| **Sero +** | **48 (96)** | **65 (87)** |
| *PCR- sero+ MLF+* | *3 (6)* | *9 (12)* |
| *PCR- sero+ MLF-* | *0* | *13 (17)* |
| **MLF +** | **46 (92)** | **57 (76)** |
| *PCR- sero- MLF+* | *1 (2)* | *5 (7)* |

**Table S2 | PCR, serum and mucosal positivity against SARS-CoV-2 in index and household cases.** The PCR positive threshold was set at a Ct value <36, corresponding to a viral load of $\geq 10^3$ copies/ml of extracted sample. The seroconversion threshold was based on the mean + 2*SD of the log2-transformed pre-SARS-CoV-2 control samples. To identify cases based on mucosal antibodies against S, a naive Bayes model was used, trained by the mucosal antibody measurements of the PCR positive study participants (cases) and the pre-SARS-CoV-2 control MLF samples (controls).
