## Supplementary material for "Elevated mucosal antibody responses against SARS-CoV-2 are correlated with lower viral load and faster decrease in systemic COVID-19 symptoms": Figure S1

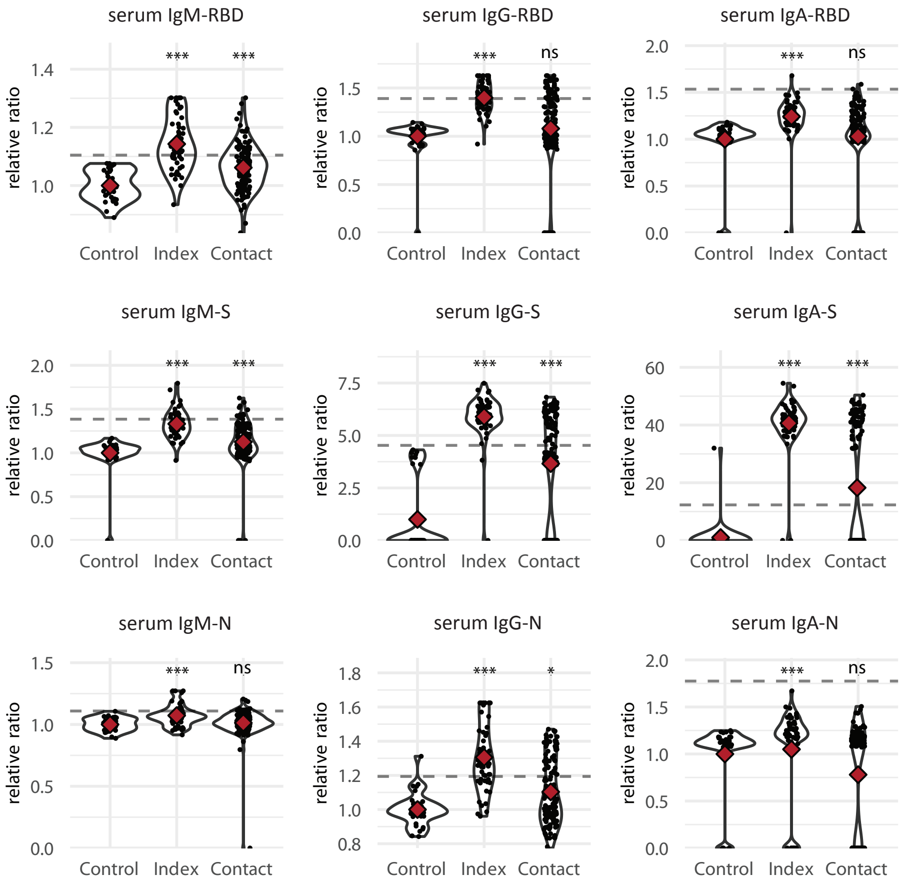


**Figure S1. Serum antibody responses against Sars-CoV-2 in COVID-19 patients and household contacts.** IgM, IgG, and IgA serum antibody responses against Sars-CoV-2 spike protein (S), receptor binding domain (RBD), or nucleocapsid (N). Data are shown for pre-SARS-Cov-2 control samples (N=32), index cases (N=50), or household contacts (N=129). Antibody levels are expressed as a ratio compared to the mean of the controls (relative ratio) and values of indexes and contacts were compared to those of controls using a Wilcoxon signed rank test, * p <0.05; ** p<0.01; *** p<0.001, and a threshold for seroconversion (dotted line) was calculated based on the mean + 2*sd of the control samples.
