## Supplementary material for "Elevated mucosal antibody responses against SARS-CoV-2 are correlated with lower viral load and faster decrease in systemic COVID-19 symptoms": Figure S4

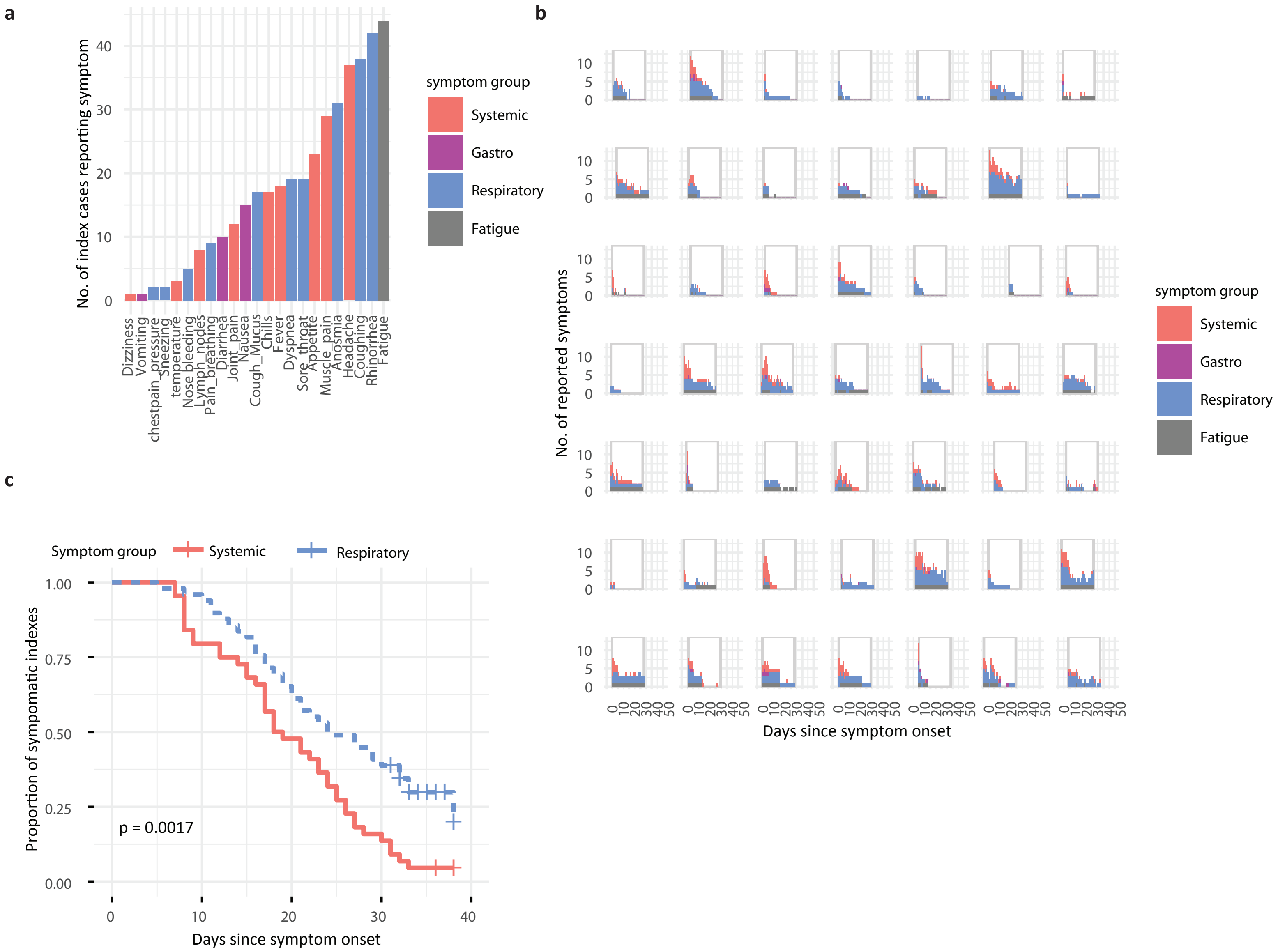


**Figure S4. Clinical presentation of mild COVID-19 disease. a)** Index cases (N = 49) completed a daily symptom survey covering 23 symptoms for 28 days during the study period. The number of index cases that reported a given symptom at any time during the study period is presented. Symptoms were categorized into either respiratory symptoms (RS), systemic disease symptoms (SDS), or gastrointestinal symptoms (GS). Fatigue was not categorized. **b)** Cumulative symptoms per symptom group for each study day are plotted for each index case. Data are represented since the time of symptom onset. A window depicts the study period for a given index case. **c)** Symptom durations were calculated for RS and SDS, and compared using a Kaplan Meier analysis. The probability of becoming symptom-free at a given time post-symptom onset is depicted. The log-rank p-value represents the statistical difference between resolution of SDS and RS.
