## Supplementary material for "Elevated mucosal antibody responses against SARS-CoV-2 are correlated with lower viral load and faster decrease in systemic COVID-19 symptoms": Figure S5

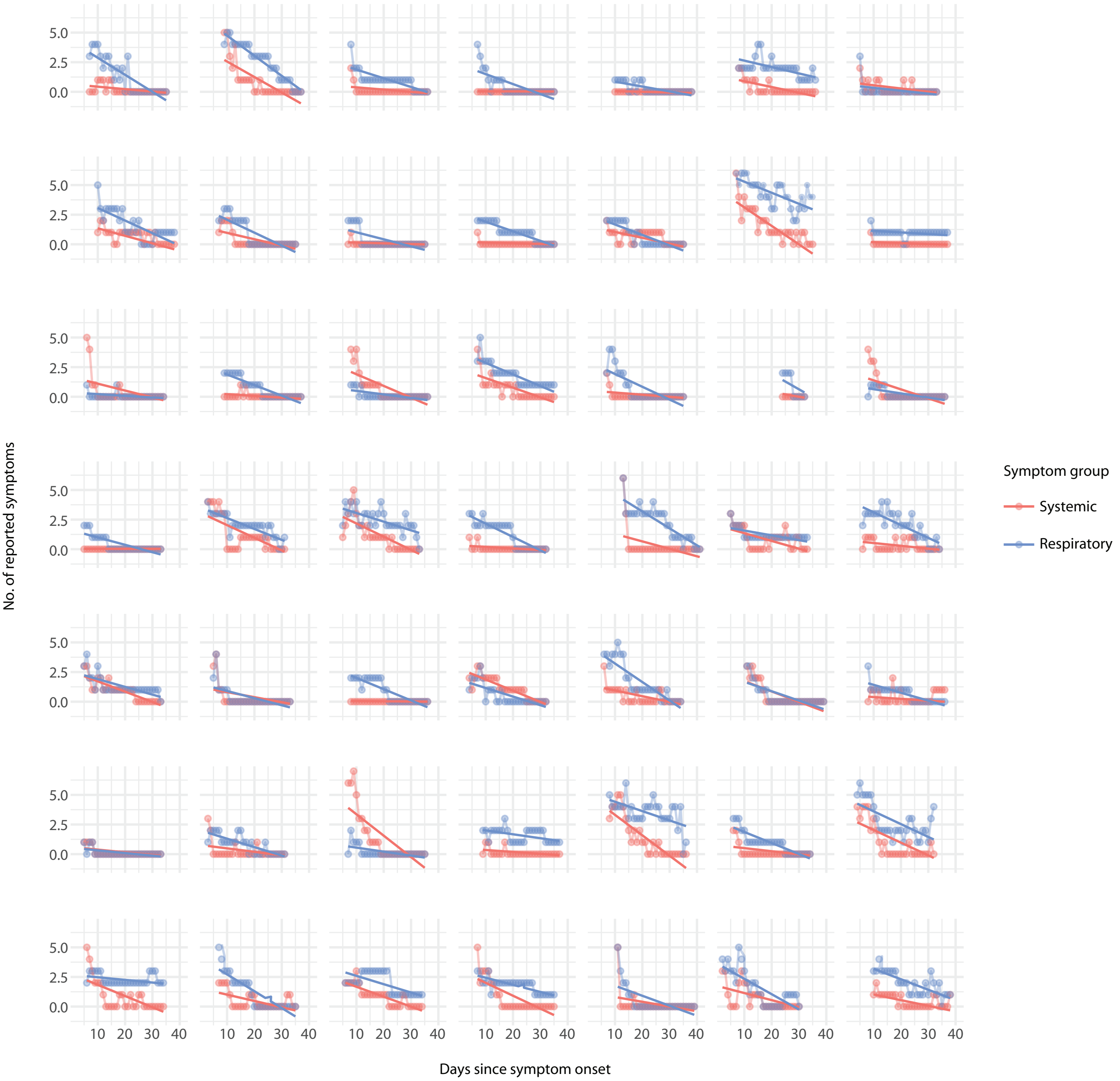


**Figure S5. Mixed-effects modelling of longitudinal symptom data in COVID-19 patients.** The number of respiratory (RS) and systemic disease (SDS) symptoms were determined for index cases (N = 49) for each day during the 28 day study period. Data are represented relative to the time of symptom onset. A linear mixed-effect model was fit to the data per symptom group. The response was specified as the number of symptoms on a given day, and explanatory fixed effects variables were: day since symptom onset, age, and sex. Time since symptom onset was also specified as a random slope, and Sample ID as a random intercept. Datapoints are plotted per day and joined by a faded line, predicted values from the mixed-effects model are plotted on top as solid straight lines.
