## Supplementary material for "Elevated mucosal antibody responses against SARS-CoV-2 are correlated with lower viral load and faster decrease in systemic COVID-19 symptoms": Figure S6

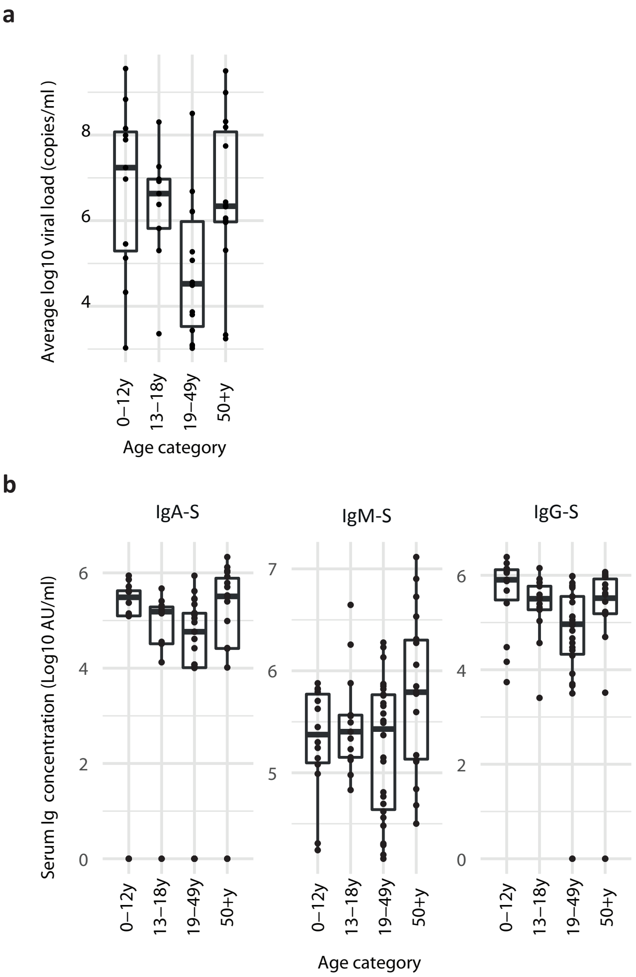


**Figure S6. Viral load and anti-Spike serum antibody concentrations in household contact cases of different age categories. a)** Viral load at study enrolment, depicted for the PCR positive household cases (n=46). The y-axis shows the average viral load in log10 copies/ml extracted sample of the contact cases, the x-axis shows the four different age categories. **b)** Serum anti-Spike protein levels at the end of the study, for IgA, IgM and IgG, in each age category. Serum antibody levels are depicted as the log10 antibody concentration in AU/ml. Only infected household members were selected (n=75).
