## Supplementary material for "Elevated mucosal antibody responses against SARS-CoV-2 are correlated with lower viral load and faster decrease in systemic COVID-19 symptoms": Figure S2

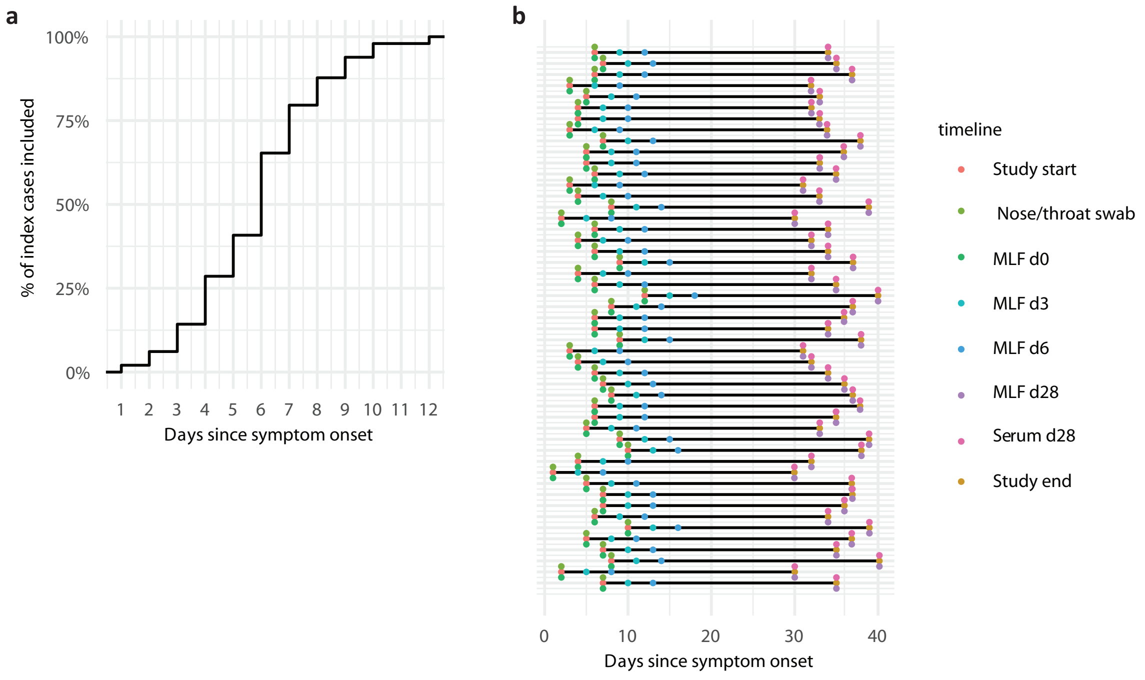


**Figure S2. Timing of COVID-19 patient recruitment and sample measurements relative to symptoms onset. a)** The graph shows the cumulative percentage of households (N=50) included at a given time post symptom onset. **b)** Overview of the measurements made for the index cases relative to the reported first day of symptom onset. Each line represents one participant.
