## Supplementary material for "Elevated mucosal antibody responses against SARS-CoV-2 are correlated with lower viral load and faster decrease in systemic COVID-19 symptoms": Figure S3

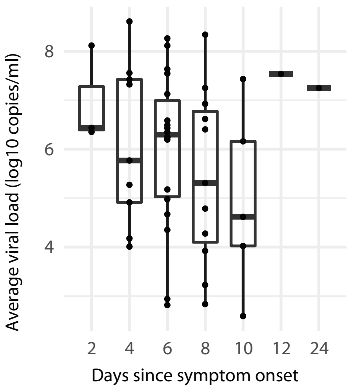


**Figure S3. SARS-CoV-2 viral load at different days post symptom onset.** Index cases (N=49) were asked when their symptoms started, and viral load was measured at study start. Individuals are binned together based on their reported symptom onset, relative to the study start. The y-axis shows the average viral load in log10 copies/ml extracted sample.
